# The Circadian Disruption Index: development, validation, and responsiveness to circadian health education

**DOI:** 10.64898/2026.07.08.26357517

**Authors:** Yimei Fan, Mingyu Tian, Jiaying Xu, Mingxin Cao, Nana Zheng, Yaping Liu, Ai, Yannis Yan Liang, Jing Wang, Xiaoqing Hu, Xiao Tan, Christian Benedict, Yun Kwok Wing, Jihui Zhang, Hongliang Feng

## Abstract

**Study Objectives:** To develop and initially validate the Circadian Disruption Index (CDI), a self-report measure of circadian disruption, and obtain preliminary evidence of its responsiveness to circadian health education.

**Methods:** In Study 1, 244 participants completed a 22-item CDI version and external measures. The sample was randomly divided for exploratory and confirmatory factor analyses. Internal consistency, external associations, and discrimination of poor sleep quality were examined. In Study 2, 72 postgraduate students completed the CDI before and 1 week after a 16-hour circadian health education program in an uncontrolled pre-post design.

**Results:** Analyses yielded a 15-item, three-factor structure comprising rhythm stability and light exposure, behavioral habits and diet, and sleep quality and subjective complaints. Total-score internal consistency was acceptable (Cronbach’s α = 0.871). Confirmatory factor analysis showed a comparative fit index of 0.902 and a root mean square error of approximation of 0.072, although the Tucker-Lewis index was 0.882. CDI scores correlated with sleep quality, chronotype, corrected midsleep on free days, depression, and anxiety, but not social jetlag. The area under the curve for poor sleep quality was 0.807 (95% confidence interval, 0.753-0.862), with an exploratory cutoff of ≥ 23. In Study 2, CDI scores decreased from 22.26 to 19.88 (*p* = 0.002; Cohen’s *d_z_* = 0.36).

**Conclusions:** The CDI demonstrated satisfactory internal consistency, a meaningful multidimensional structure, and responsiveness to short-term changes following circadian health education, supporting its potential utility for assessing circadian disruption and monitoring circadian-related behavioral changes.

**Statement of Significance:** Existing questionnaires typically assess isolated aspects of circadian functioning, such as chronotype, sleep timing, or sleep quality, leaving a need for a brief measure that captures the broader burden of circadian disruption. This study introduces the Circadian Disruption Index, which integrates rhythm stability and light exposure, behavioral habits and diet, and sleep-related complaints within a single self-report instrument. Initial findings support its multidimensional structure and short-term responsiveness following circadian health education. The index may provide a practical tool for population research and the evaluation of circadian-focused interventions.

## Introduction

Circadian rhythms are endogenous, approximately 24-hour biological timing systems that regulate a wide range of physiological and behavioral processes (e.g., the sleep–wake cycle, endocrine secretion, metabolism, cognitive performance, and mood regulation) [1]. In modern society, individuals are increasingly exposed to a variety of circadian risk factors, including excessive use of electronic devices before bedtime, shift work, nighttime light exposure, and insufficient daytime light exposure [2, 3]. As a result, approximately 60% of the population experience circadian disruption, such as delayed sleep, and social jetlag (SJL) [4, 5]. Circadian disruption has been associated with widespread adverse physiological and psychological consequences and increases the risks of traffic accidents and occupational errors [6–9]. Accordingly, feasible and scalable approaches to assessing circadian disruption are needed for epidemiological research, risk assessment, and the evaluation of behavioral interventions.

To date, non-invasive, low-burden, and scalable tools for quantifying circadian disruption in large clinical and epidemiological samples remain limited [10, 11]. Dim light melatonin onset (DLMO), widely regarded as the gold-standard marker of human circadian timing, and core body temperature monitoring are reliable methods for assessing circadian rhythms [1, 12]. However, their complex sampling procedures, high costs, and limited feasibility substantially restrict their application [13, 14]. Although actigraphy conducted over 7–14 days or longer can objectively and conveniently characterize rest–activity rhythms, heterogeneity in devices, sensors, and analytical approaches has limited the development of standardized metrics for the comprehensive assessment of circadian disruption [15]. Moreover, actigraphy-derived circadian metrics are primarily based on patterns of movement and inactivity and therefore provide limited information on the environmental and other daily behavioral factors [16]. To the best of our knowledge, widely used self-report instruments for assessing circadian characteristics, such as the Morningness–Eveningness Questionnaire (MEQ) and the Munich Chronotype Questionnaire (MCTQ), primarily capture diurnal preference or habitual sleep timing and SJL, and therefore do not fully capture the multidimensional nature of circadian disruption [17, 18].

Therefore, this study aimed to develop and validate a multidimensional self-report instrument designed to systematically quantify the severity of circadian disruption (Study 1). To develop the initial item pool, items were adapted from existing circadian-related instruments and informed by relevant theoretical and empirical evidence. The items covered major manifestations of circadian disruption, including altered sleep phase, irregular daily routines, sleep difficulties, poor sleep quality, and daytime dysfunction, as well as behavioral and environmental determinants such as meal timing, physical activity, and daytime and nighttime light exposure [19]. The item pool was refined through expert consultation to ensure content validity, conceptual relevance, and clarity before empirical testing. To assess the applicability and responsiveness of the CDI, we administered an online survey to postgraduate students participating in circadian health education and examined whether CDI scores reflected short-term changes following the education (Study 2), thereby providing preliminary evidence of the CDI’s responsiveness in this context.

## Method

### Study 1: Development and psychometric validation of the Circadian Disruption Index Study population

Participants were recruited through online advertisements and completed the survey electronically. The questionnaires were administered in Chinese. Individuals were eligible if they were aged 12 years or older, possessed sufficient reading ability to understand the questionnaire, and provided informed consent prior to participation. Although the original eligibility criterion permitted participation from the age of 12 years, all participants included in the present analysis were aged 18 years or older. A total of 247 questionnaires were collected. Three questionnaires were excluded because of illogical response patterns or duplicate responses, resulting in a final sample of 244 participants (Figure 1A). The total sample was randomly divided into two independent subsamples for exploratory factor analysis (EFA) and confirmatory factor analysis (CFA). The study protocol was approved by the Ethics Committee of the Affiliated Brain Hospital of Guangzhou Medical University, and all participants provided informed consent before participation.

**Figure 1.**
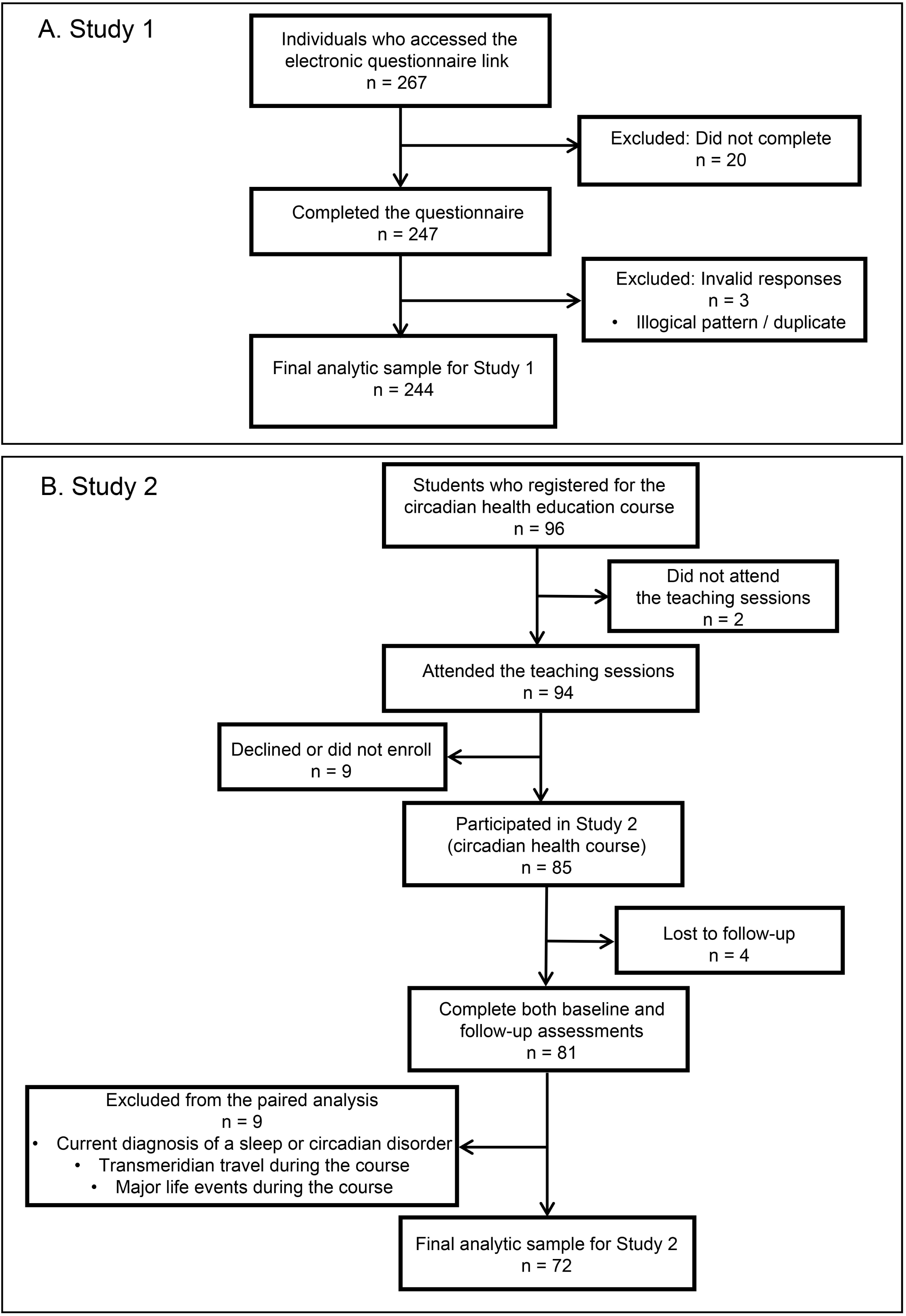
Flow diagram of participant recruitment and inclusion in Studies 1 and 2. (A) Recruitment and inclusion of participants in the CDI development and psychometric validation study. (B) Recruitment, follow-up, and inclusion of participants in the circadian health education study. Alt text: Two-panel participant flow diagram. Study 1 included 244 valid participants who were divided equally into exploratory and confirmatory factor-analysis samples. Study 2 included 72 participants in the final paired pre–post analysis.

### Measure development

CDI was developed to provide a multidimensional assessment of circadian disruption in the general population. Item generation was informed by current theoretical frameworks of circadian health, clinical characteristics of circadian rhythm sleep–wake disorders, and empirical evidence regarding behavioral and environmental factors associated with circadian disruption. An initial pool of 25 scored items was generated to assess four hypothesized domains: (1) sleep quality and daytime functioning, (2) circadian rhythm disturbance–related symptoms, (3) behavioral and feeding rhythms, and (4) light exposure and sleep environment. To establish content validity, the preliminary item pool was reviewed by an expert panel consisting of three sleep medicine specialists and two psychologists. The experts evaluated each item for relevance, clarity, and comprehensibility, and provided recommendations for revision.

Subsequently, a pilot study was conducted to evaluate the comprehensibility and acceptability of the questionnaire. Based on feedback from both the expert panel and pilot participants, three items were removed because of content overlap or insufficient clarity, resulting in a 22-item survey version for formal psychometric evaluation. In addition to the scored items, seven descriptive items were included to collect information on participants’ sleep habits and circadian characteristics, including current shift-work status, workday or school-day and free-day sleep schedules, schedules, sleep latency, nap duration, perceived sleep need, and chronotype preference. These descriptive items were not included in the calculation of the CDI total score. The 22-item survey version was rated on a five-point Likert scale ranging from 0 (“never”) to 4 (“always”), with higher scores indicating greater circadian disruption. The total CDI score was calculated by summing the scores of all 22 items. The 22-item CDI is seen in Supplementary Appendix S1.

### Validation Measures

To evaluate criterion-related validity evidence for the CDI, participants completed several validated questionnaires. Sleep quality was assessed using the Pittsburgh Sleep Quality Index (PSQI), with higher scores indicating poorer sleep quality [20]. A global PSQI score >5 is commonly used to identify poor sleep quality. Chronotype was evaluated using the reduced version of the MEQ, where higher scores indicated greater morningness [17, 21]. Circadian timing variables, including social jetlag (SJL) and corrected midsleep on free days (MSFsc), were derived from the MCTQ [22]. Depressive and anxiety symptoms were evaluated using the Patient Health Questionnaire-9 (PHQ-9) and the Generalized Anxiety Disorder-7 (GAD-7) [23, 24]. These measures were selected to examine the relationships between CDI scores and established indicators of sleep quality, circadian timing, chronotype, and psychological functioning.

### Statistical analysis

All statistical analyses were conducted using Python (version 3.11.15). Continuous variables were summarized as means and standard deviations, and categorical variables as frequencies and percentages. The pingouin package was used for reliability and correlation analyses, factor_analyzer for item analysis and exploratory factor analysis, semopy for confirmatory factor analysis, and scikit-learn for receiver operating characteristic (ROC) analyses.

Item analysis was performed for the initial 22-item CDI. Participants were ranked according to their total CDI scores, and those in the upper and lower 27% of the score distribution were classified as the high- and low-score groups, respectively. Item discrimination was evaluated by comparing item scores between the two groups using independent-samples t tests, with the resulting t statistic reported as the critical ratio (CR). A CR value > 3.00 with *p* < 0.05 was considered indicative of acceptable item discrimination [25]. Corrected item-total correlations were examined to assess item homogeneity, with values of 0.40 or higher indicating adequate item internal consistency [26]. Cronbach’s alpha if item deleted was also considered when evaluating item retention. Items were retained or removed based on a combined assessment of discrimination, item-total correlation, internal consistency, factor structure, and item interpretability.

The total sample was randomly divided in a 1:1 ratio into Sample A for EFA and Sample B for CFA using a computer-generated random sequence. In Sample A (n = 122), the suitability of the data for EFA was assessed using the Kaiser-Meyer-Olkin (KMO) measure and Bartlett’s test of sphericity. A KMO value above 0.80 was considered good, and a significant Bartlett’s test indicated that the correlation matrix was appropriate for factor analysis [27]. Factors were extracted using the minimum residual method and subjected to Promax oblique rotation because the underlying dimensions were expected to be correlated. The number of factors retained was determined by considering eigenvalues greater than 1.0, visual inspection of the scree plot, and the conceptual interpretability of the resulting factor solution. Items with factor loadings of 0.40 or higher were considered acceptable for retention [27].

In Sample B (n = 122), CFA was used to evaluate the factor structure identified by EFA. Model fit was assessed using the chi-square to degrees-of-freedom ratio (χ²/df), comparative fit index (CFI), Tucker-Lewis index (TLI), root mean square error of approximation (RMSEA), and standardized root mean square residual (SRMR). A χ²/df below 3, CFI and TLI values above 0.90, and RMSEA and SRMR values below 0.08 were considered indicative of acceptable model fit [28]. Standardized factor loadings of 0.40 or higher were considered interpretable [29].

Internal consistency was assessed using Cronbach’s alpha. Composite reliability and average variance extracted (AVE) were calculated to further evaluate the reliability and convergent validity of each factor. Cronbach’s alpha and composite reliability values of 0.70 or higher were considered acceptable, whereas AVE values of 0.50 or higher were considered evidence of adequate convergent validity [29]. Criterion-related validity was examined using Pearson correlation analyses between CDI scores and external measures, including PSQI, rMEQ, MCTQ-derived SJL and MSFsc, PHQ-9, and GAD-7.

Additional hypothesis-based construct validity analyses examined associations between CDI scores and age, and compared CDI scores across sex, usual daytime work/school and nighttime sleep schedule, and chronotype groups. Chronotype trends were further evaluated using Spearman correlation. ROC analysis was conducted in the full Study 1 sample to evaluate the ability of the CDI total score to identify poor sleep quality, defined as a PSQI global score >5. The area under the ROC curve (AUC) and its 95% confidence interval were calculated, and the optimal cutoff was selected by maximizing the Youden index. Sensitivity and specificity were calculated at the selected cutoff. Statistical significance was defined as a two-sided *p* < 0.05.

### Study 2: Responsiveness to circadian health education Study population

Participants were postgraduate students from Guangzhou Medical University enrolled in circadian health education in January 2026. A total of 96 students registered for it, and 94 attended the teaching sessions. Questionnaires were administered online before circadian health education and 1 week after its completion. All participants provided informed consent before completing the baseline questionnaires. A total of 85 participants submitted valid baseline questionnaires, of whom four did not complete the follow-up assessment. Among the remaining 81 participants with paired questionnaires, nine were excluded because they reported a current sleep- or circadian rhythm-related disorder, trans-time-zone travel during teaching sessions, or a major life event during the period. The final paired analytic sample included 72 participants (Figure 1B).

### The circadian health education

Circadian health education was a brief, intensive educational program comprising 16 teaching hours delivered over four consecutive days, from January 16 to 19, 2026. Sessions were held daily from 17:50 to 20:40. Participants were reassessed 1 week after completion to evaluate short-term responsiveness to the education. The content covered the basic principles and mechanisms of circadian rhythms, circadian assessment methods, clinical regulation strategies, circadian rhythm-related sleep disorders, circadian disruption in neuropsychiatric and cardiometabolic diseases, systemic applications, and modern circadian challenges such as shift work, jet lag, and nocturnal light exposure.

### Assessment of responsiveness to circadian health education

The CDI was the primary responsiveness measure. The final CDI contains 15 scored items rated from 0 (“never”) to 4 (“always”), with higher scores indicating greater circadian disruption. For comparison, the PSQI, rMEQ, MCTQ, PHQ-9, and GAD-7 were also assessed, as in Study 1. In addition, the Epworth Sleepiness Scale (ESS) and Stanford Presenteeism Scale-6 (SPS-6) were administered to reflect daytime functioning during study activities. The PSQI was used for baseline sleep-quality characterization only. MCTQ-derived timing variables included workday and free-day sleep onset, wake time, sleep duration, midsleep, MSFsc, and SJL.

### Statistical analysis

Analyses were conducted using Python version 3.11.15. Paired pre-post comparisons used two-sided Wilcoxon signed-rank tests on raw scores. All hypothesis tests were conducted using the measures in their original scoring directions. Effect-size signs were harmonized only for comparative visualization. Paired Cohen’s *d_z_*was then calculated separately, with the direction harmonized so that positive values indicated improvement across measures. Item-level response distributions were summarized as category percentages; category-level changes were tested using two-sided exact McNemar tests, and item-score changes using Wilcoxon signed-rank tests. Item-and category-level *p* values were considered exploratory and were not adjusted for multiple comparisons. Analyses used complete paired observations without imputation. Statistical significance was defined as two-sided *p* < 0.05.

## Results

### Study 1: Development and psychometric validation of the Circadian Disruption Index Item analysis

A total of 244 participants with valid responses were included in the final analysis. Their mean age was 23.19 ± 5.96 years, with an age range of 18–50 years, and 141 participants (57.8%) were women (Table 1). Item analysis was conducted using critical ratio tests, corrected item-total correlations, and internal consistency analysis. The initial 22-item CDI demonstrated good internal consistency, with an overall Cronbach’s alpha of 0.887. CR values ranged from 3.09 to 12.84, and all comparisons between the high- and low-score groups were statistically significant (all *p* < 0.01). Corrected item-total correlations ranged from 0.13 to 0.68. Specifically, Q12, Q14, Q16, Q17, Q19, Q27, and Q28 were excluded because their item-total correlations were below the prespecified threshold of 0.40. After integrating empirical item performance with conceptual relevance, 15 core items were retained for subsequent factor analysis. The detailed results are shown in Table 2.

### Exploratory factor analysis

Exploratory factor analysis was performed on the refined 15-item CDI using Sample A. The KMO value was 0.847, and Bartlett’s test of sphericity was significant (χ² = 701.29, *p* < 0.001). A three-factor solution with eigenvalues greater than 1 was retained (Figure 2). The eigenvalues of the three factors were 5.71, 1.50, and 1.17, accounting for 34.77%, 6.99%, and 4.07% of the total variance, respectively. The cumulative variance explained by the three-factor solution was 45.83%.

**Figure 2.**
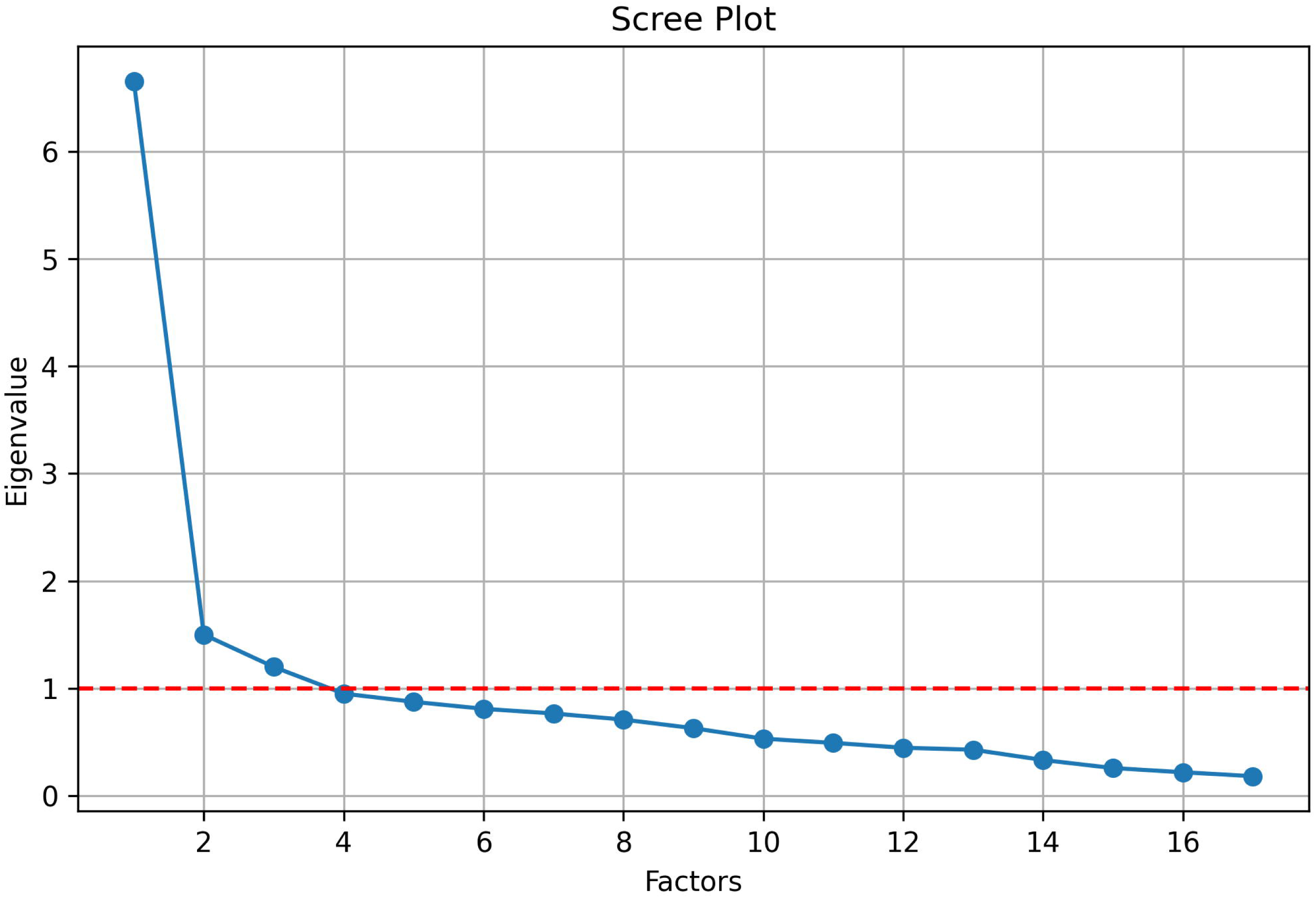
Scree plot for factor retention in the exploratory factor analysis of the Circadian Disruption Index. The first three factors had eigenvalues greater than 1.0, followed by a flattening of the curve, supporting the retention of a three-factor solution. The red dashed line represents the Kaiser criterion of an eigenvalue equal to 1.0. Alt text: Scree plot showing that the first three factors had eigenvalues greater than 1, followed by a flattening of the curve.

Following Promax rotation, five items loaded on each factor. Factor 1 comprised Q13, Q15, Q21, Q25, and Q26 and was labeled rhythm stability and light exposure, with factor loadings ranging from 0.47 to 0.92. Factor 2 comprised Q18, Q20, Q22, Q23, and Q24 and was labeled behavioral habits and diet, with loadings ranging from 0.49 to 0.78. Factor 3 comprised Q8, Q9, Q10, Q11, and Q29 and was labeled sleep quality and subjective complaints, with loadings ranging from 0.50 to 0.92. All retained items had primary factor loadings greater than 0.40 (Table 3).

### Confirmatory factor analysis

Confirmatory factor analysis was conducted using Sample B to evaluate the three-factor structure identified in the exploratory factor analysis. The three-factor model yielded a χ²/df ratio of 1.63, a CFI of 0.902, a TLI of 0.882, an RMSEA of 0.072, and an SRMR of 0.077. Overall, the model demonstrated acceptable fit, although the TLI was slightly below the prespecified threshold of 0.90. All 15 items loaded significantly on their respective latent factors, with standardized factor loadings ranging from 0.410 to 0.793 (all *p* < 0.001).

### Reliability, criterion-related validity, and discriminatory performance

The Cronbach’s α coefficients were 0.747, 0.729, and 0.808 for F1, F2, and F3, respectively, while the Cronbach’s α coefficient for the CDI total score was 0.871. Composite reliability values were 0.735, 0.732, and 0.821, respectively, and all exceeded the prespecified threshold of 0.70. AVE values were 0.364, 0.363, and 0.480 for F1, F2, and F3, respectively, and therefore did not reach the prespecified threshold of 0.50 (Table 4).

The three factor scores were significantly correlated with the CDI total score, with correlation coefficients ranging from 0.777 to 0.865 (all *p* < 0.001). Correlations among the three factors ranged from 0.457 to 0.585. The CDI total score was positively correlated with PSQI (r = 0.664, *p* < 0.001), MSFsc (r = 0.289, *p* < 0.001), PHQ-9 (r = 0.486, *p* < 0.001), and GAD-7 (r = 0.291, *p* < 0.001), and negatively correlated with rMEQ (r = −0.192, *p* < 0.01). The correlation between the CDI total score and SJL was not significant. Among the three factors, F3 showed the strongest correlation with PSQI (r = 0.722, *p* < 0.001) (Table 5).

Additional analyses examining hypothesis-based construct validity showed that CDI total scores were not significantly associated with age and did not differ significantly by sex or usual daytime work/school and nighttime sleep schedule. In contrast, CDI scores differed significantly across the five chronotype groups (Kruskal–Wallis *p* < 0.001) and showed a significant monotonic association with increasing eveningness (Spearman’s ρ = 0.262, *p* < 0.0001), with definite evening types exhibiting higher CDI scores than moderate morning and intermediate types after Holm adjustment. ROC analysis was performed to evaluate the ability of the CDI total score to identify poor sleep quality defined by the PSQI. Using PSQI > 5 as the criterion for poor sleep quality, the CDI total score yielded an AUC of 0.807 (95% CI, 0.753–0.862). The optimal cutoff identified using the Youden index was a CDI total score of ≥ 23, with a sensitivity of 73.6% and a specificity of 78.0% (Figure 3).

**Figure 3.**
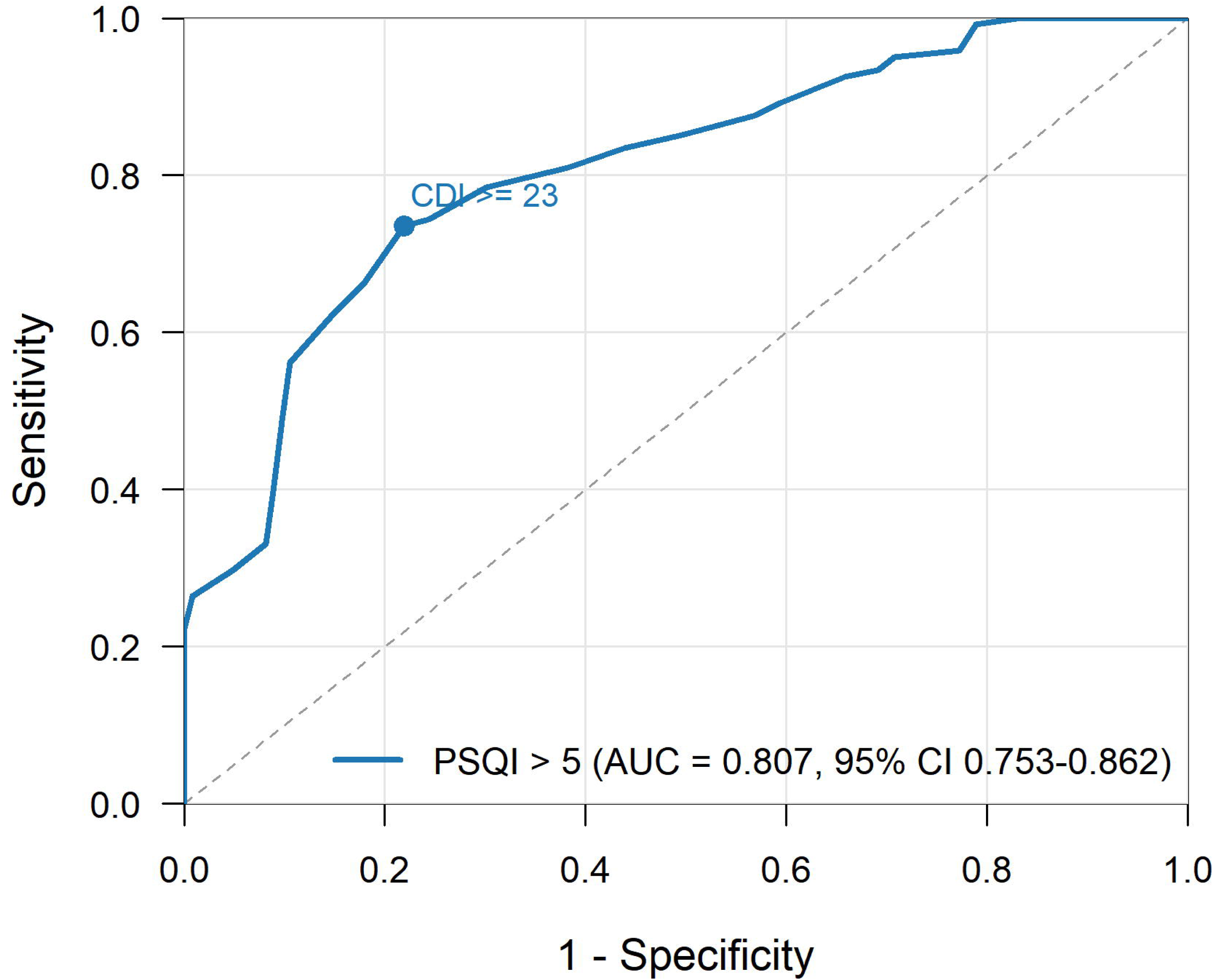
Receiver operating characteristic curve of the Circadian Disruption Index total score for identifying poor sleep quality. The area under the curve (AUC) was 0.807 (95% confidence interval: 0.753–0.862). The optimal cutoff determined using the Youden index was a Circadian Disruption Index total score of ≥ 23, yielding a sensitivity of 73.6% and a specificity of 78.0%. The diagonal dashed line represents chance-level discrimination. Alt text: Receiver operating characteristic curve showing an area under the curve of 0.807 for identifying poor sleep quality using the Circadian Disruption Index total score.

### Study 2: Responsiveness to circadian health education Baseline characteristics of the analytic sample

A total of 72 participants were included in the paired analysis (Table 6). The mean age was 24.52 ± 2.37 years, and the sample included 35 women (48.6%) and 37 men (51.4%). At baseline, 35 participants (48.6%) had poor sleep quality as defined by PSQI >5. Chronotype distribution included 18 morning-type participants (25.0%), 25 intermediate-type participants (34.7%), and 29 evening-type participants (40.3%). The baseline CDI total score was 22.26 ± 8.57, with mean factor scores of 8.50 ± 3.75 for F1, 5.71 ± 2.84 for behavioral habits and diet, and 8.06 ± 3.85 for sleep quality and subjective complaints.

### Pre-post changes in sleep timing, CDI, and established measures

To provide a behavioral context for the observed changes in CDI scores, we examined sleep-timing variables derived from the MCTQ among the 72 participants with valid paired MCTQ data (Table S1). Between the two assessments, workday sleep onset advanced from 01:02 to 00:16 (*p* < 0.001), and free-day sleep onset advanced from 01:36 to 00:55 (*p* < 0.001). Workday sleep duration increased from 7.09 to 7.80 h (*p* < 0.001), and free-day sleep duration increased from 7.93 to 8.55 h (*p* = 0.001). MSFsc advanced from 05:08 to 04:46 (*p* = 0.002). In contrast, SJL did not change significantly, increasing from 0.99 to 1.07 h (*p* = 0.505) (Figure 4). Overall, lower CDI scores at the second assessment coincided with earlier sleep timing and longer sleep duration.

**Figure 4.**
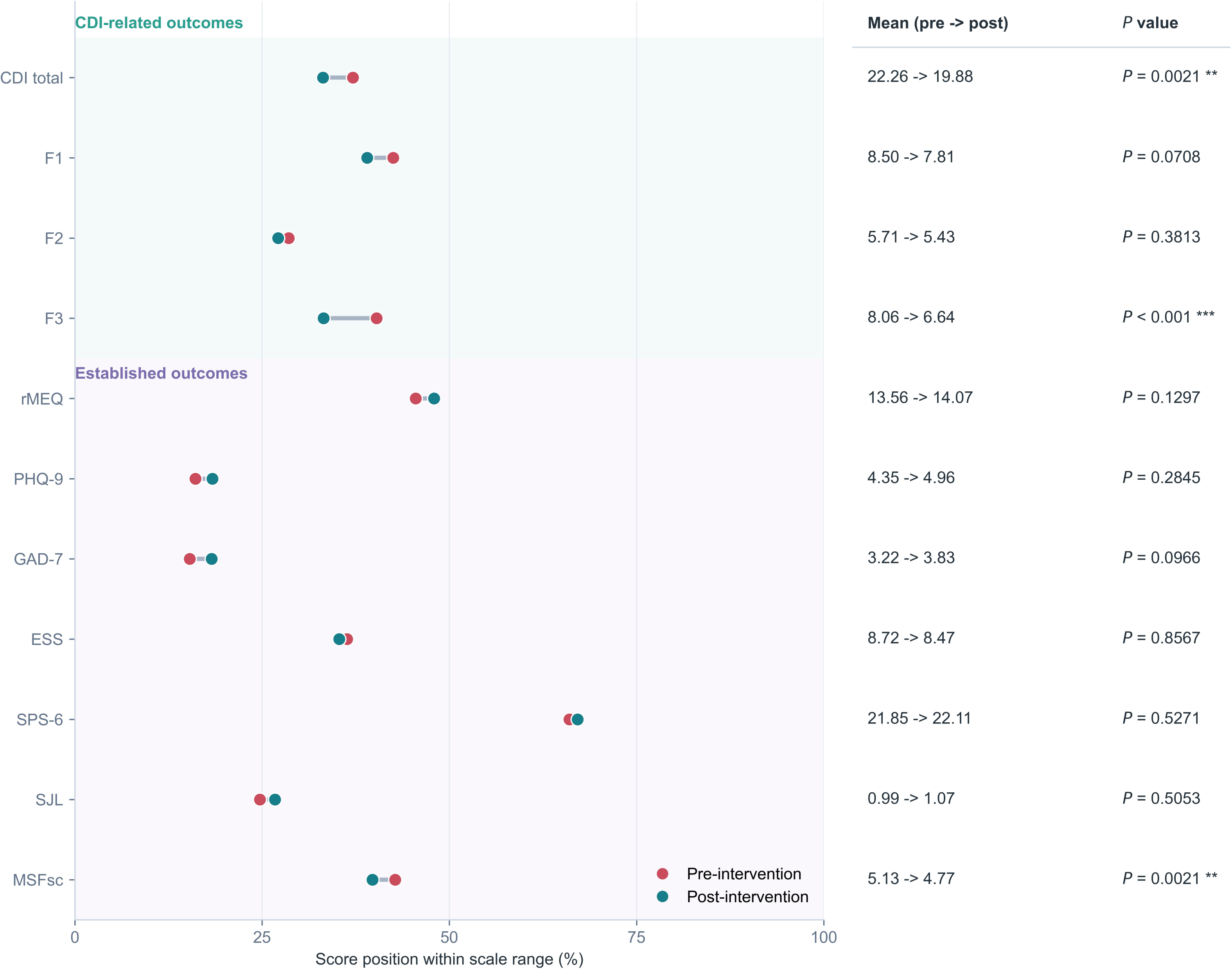
Pre- and post-Circadian Health Education scores for the CDI and established outcome measures. Dots indicate the pre- and post-intervention means, and connecting lines indicate the direction of the mean change within each outcome. Because the measures have different scoring ranges, each mean was mapped to its instrument-specific range for joint visualization; the untransformed means and two-sided Wilcoxon signed-rank *p* values are shown at right. No symbol indicates *p* ≥ 0.05; \**p* < 0.05, \*\**p* < 0.01, and \*\*\**p* < 0.001. Lower scores indicate improvement for the CDI, PHQ-9, GAD-7, ESS, and SJL; higher scores indicate greater morningness for the rMEQ and better function for the SPS-6. A lower MSFsc indicates an earlier sleep midpoint and a more morning-type circadian phenotype. CDI, Circadian Disruption Index; ESS, Epworth Sleepiness Scale; GAD-7, Generalized Anxiety Disorder-7; PHQ-9, Patient Health Questionnaire-9; rMEQ, reduced Morningness-Eveningness Questionnaire; SJL, social jetlag; MSFsc, sleep-corrected midpoint of sleep on free days; SPS-6, Stanford Presenteeism Scale-6. SJL and MSFsc are reported in hours. Alt text: Paired pre- and post-education means are shown for the CDI and comparator measures. The largest visible reductions occurred in the CDI total score and the sleep quality and subjective complaints factor, while MSFsc shifted to an earlier time and most other measures changed little.

The CDI total score decreased from 22.26 ± 8.57 at baseline to 19.88 ± 9.57 at follow-up, corresponding to a mean change of −2.39 (*p* = 0.002) (Figure 4). Among the three CDI factors, the largest change was observed for F3, which decreased from 8.06 ± 3.85 to 6.64 ± 3.85 (*p* < 0.001). F1 and F2 showed a smaller reduction but did not reach statistical significance. MSFsc advanced significantly, whereas the other comparator measures showed no significant pre–post changes. Consistent with the factor-level findings, the supplementary item-level analysis showed that significant item-score reductions were concentrated in late bedtime, sleep-related complaints, snacking outside regular meals, and sleep-environment sensitivity (Figure S1).

### Standardized effect sizes across outcomes

The direction-harmonized effect-size analysis further indicated that the short-term response was concentrated in CDI-related outcomes (Figure 5). The largest standardized change was observed for sleep quality and subjective complaints (Cohen’s *d_z_* = 0.53), followed by MSFsc (Cohen’s *d_z_* = 0.38) and the CDI total score (Cohen’s *d_z_*= 0.36). Rhythm stability and light exposure showed a small-to-moderate effect (Cohen’s *d_z_* = 0.22), while behavioral habits and diet showed only a small effect (Cohen’s *d_z_* = 0.10). Other comparator measures showed small and mixed effects, including rMEQ (Cohen’s *d_z_* = 0.21), ESS (Cohen’s *d_z_* = 0.07), SPS-6 (Cohen’s *d_z_* = 0.08), SJL (Cohen’s *d_z_* = -0.10), PHQ-9 (Cohen’s *d_z_* = -0.16), and GAD-7 (Cohen’s *d_z_* = -0.22).

**Figure 5.**
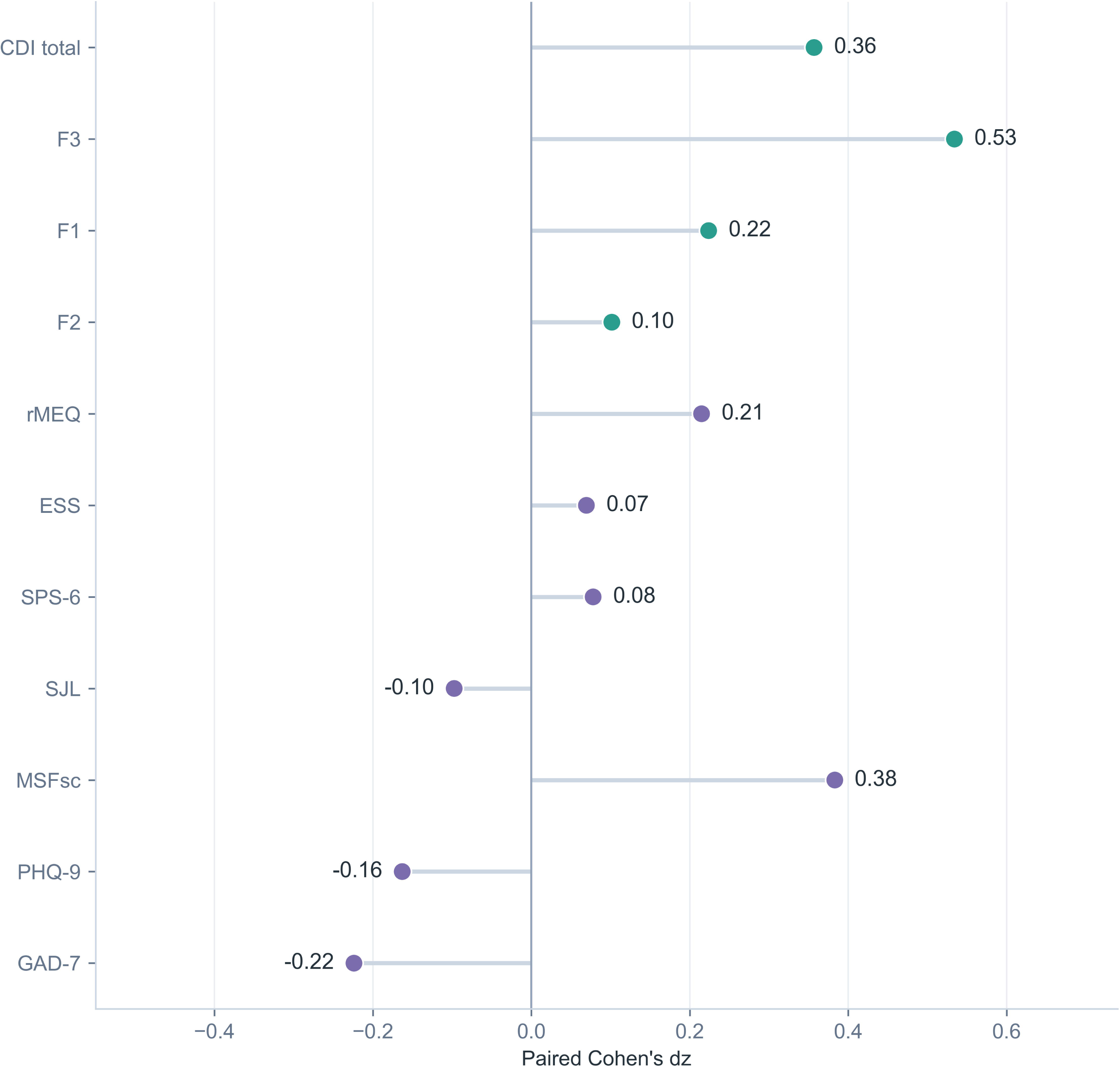
Direction-harmonized paired Cohen’s *d_z_* effect sizes for the CDI and established outcome measures. Points indicate paired Cohen’s *d_z_* values, calculated from the participant-level change scores, and horizontal lines extend from the null value of zero. Effect-size direction was harmonized so that positive values indicate improvement or change in the target direction. Teal points denote CDI-related outcomes, and purple points denote established outcome measures. CDI, Circadian Disruption Index; ESS, Epworth Sleepiness Scale; GAD-7, Generalized Anxiety Disorder-7; PHQ-9, Patient Health Questionnaire-9; rMEQ, reduced Morningness-Eveningness Questionnaire; SJL, social jetlag; MSFsc, sleep-corrected midpoint of sleep on free days; SPS-6, Stanford Presenteeism Scale-6. SJL and MSFsc are reported in hours. Alt text: Standardized paired effect sizes are displayed with positive values indicating improvement. The largest positive effect was observed for sleep quality and subjective complaints, followed by MSFsc and the CDI total score, whereas the remaining measures showed small or negative effects.

These findings indicate that in addition to specific sleep timing variables (e.g., MSFsc), the CDI captured a measurable short-term change after circadian health education, whereas conventional symptom and functional measures showed limited change over the same period.

## Discussion

The present study developed and validated the CDI, a brief multidimensional instrument designed to assess circadian disruption. Through a systematic scale-development process, a stable three-factor structure comprising rhythm stability and light exposure, behavioral habits and diet, sleep quality and subjective complaints was identified. The final version of the index is provided as a separate supplementary file (Supplementary Appendix S2). The final instrument demonstrated satisfactory internal consistency, construct validity, and criterion-related validity. CDI scores were significantly associated with established measures of chronotype, sleep quality, circadian timing, and emotional symptoms, supporting its ability to capture multiple dimensions of circadian disruption. Furthermore, preliminary findings suggested that the CDI was responsive to circadian health education, indicating its potential utility for evaluating circadian-related intervention outcomes.

To date, no previous brief and validated self-report instrument has been designed specifically to quantify the overall severity of circadian disruption across environmental, behavioral, and sleep-related domains in the general population. Widely used instruments generally characterize specific components of circadian or sleep functioning. For example, the MEQ assesses diurnal preference [17], the MCTQ estimates chronotype and SJL from habitual sleep timing [18], the PSQI evaluates subjective sleep quality and disturbances [20], and sleep diaries prospectively document day-to-day sleep patterns [30]. Although these measures are valuable for their intended purposes, none alone provides an integrated estimate of the overall burden of circadian disruption. DLMO is widely regarded as a reference marker of endogenous circadian phase; however, its assessment requires controlled lighting conditions, repeated biological sampling, and laboratory assays, limiting its feasibility for large-scale or repeated assessments [31]. The CDI addresses this gap by integrating rhythm stability and light exposure, behavioral habits and diet, and sleep-related symptoms within a single concise framework. This multidimensional structure is consistent with the emerging view that circadian health reflects the interaction among environmental time cues, behavioral timing, and sleep processes [10]. Accordingly, the CDI is not intended to replace objective circadian phase markers, but rather to complement them by providing a scalable and interpretable estimate of multidimensional circadian disruption.

Another notable finding was the responsiveness of the CDI to circadian health education. Although educational and behavioral interventions targeting sleep and circadian health have been increasingly implemented in university and healthcare settings, evaluation has often relied on traditional sleep-related outcomes such as sleep quality or sleep duration [32]. Previous studies have reported inconsistent improvements in these measures following educational interventions [33–36]. This may partly reflect a mismatch between the immediate targets of such interventions and the outcomes used to evaluate them. Circadian health education primarily seeks to modify proximal determinants of circadian regulation, including the timing and regularity of daily activities, light exposure, sleep scheduling, and related lifestyle behaviors, whereas improvements in sleep quality or duration may occur later and may also be influenced by factors beyond circadian behavior. By incorporating several of these modifiable determinants, the CDI may capture early changes in circadian-related behavioral patterns that are not fully reflected by conventional sleep questionnaires. Consistent with this interpretation, the reduction in CDI scores was accompanied by earlier sleep onset on both workdays and free days, longer sleep duration, and an earlier MSFsc. The observed reduction in CDI scores following education therefore provides preliminary support for its potential use as an intervention outcome measure.

The findings also have several potential practical implications. As a brief and easily administered self-report index, the CDI may be suitable for large-scale epidemiological surveys and population-based screening. In clinical settings, the CDI may serve as an adjunctive assessment tool to identify individuals with a greater burden of circadian-related behavioral, environmental, and sleep disturbances, thereby informing the need for more detailed evaluation using sleep histories, sleep diaries, actigraphy, or biological circadian markers. The preliminary cutoff identified in the present study may facilitate risk stratification for poor sleep quality, although it should not be interpreted as a diagnostic threshold for circadian rhythm sleep–wake disorders. In addition, because the CDI includes potentially modifiable domains such as light exposure, daily regularity, meal timing, and sleep-related behaviors, its factor- and item-level scores may help identify specific targets for personalized health education or behavioral intervention.

Several limitations should be acknowledged. First, participants were primarily university students and young adults, which may limit the generalizability of the findings to other age groups and clinical populations. Second, all measures were self-reported and may therefore be affected by recall and reporting biases. Future studies incorporating objective circadian measures, such as actigraphy-derived rhythm parameters or melatonin timing, could provide further evidence regarding the validity of the CDI. Third, although the overall psychometric findings supported the proposed three-factor structure, several indicators, including AVE values and the TLI, did not uniformly reach their ideal benchmarks. Given that conventional cutoff values should be regarded as interpretive guidelines rather than absolute criteria, these findings should be considered alongside the broader pattern of model-fit and reliability evidence [37, 38]. Further cross-validation in larger and more heterogeneous samples is warranted to confirm the stability and generalizability of the factor structure [39]. Finally, the responsiveness of the CDI was examined over a relatively short period and without extensive longitudinal validation. Controlled studies with repeated assessments and longer follow-up are needed to determine the stability, sensitivity to change, and interpretability of intervention-related changes in CDI scores. Despite these limitations, the present study provides initial evidence for the reliability, validity, and practical utility of the CDI as a multidimensional measure of circadian disruption.

In summary, the present study developed and preliminarily validated the CDI as a brief, multidimensional self-report index for assessing circadian disruption across rhythm stability and light exposure, behavioral habits and diet, and sleep quality and subjective complaints. The CDI demonstrated satisfactory reliability, a meaningful three-factor structure, and good validity and discriminatory performance. Preliminary findings further suggested that the CDI may be responsive to short-term changes following circadian health education, particularly in sleep-related complaints and modifiable circadian behaviors. The CDI may therefore provide a practical and scalable complement to existing circadian and sleep assessments in research, health education, and population-based screening, although further validation in larger, more diverse, and longitudinal samples is required.

## Data Availability

All data produced in the present study are available upon reasonable request to the authors

## Acknowledgements

The authors thank all participants for their time and contribution to this study.

## Funding

H.F. was supported by the National Natural Science Foundation of China (82571696), the National Science and Technology Innovation 2030 of China-Major Projects (2022ZD0214100), General Program of Natural Science Foundation of Guangdong Province (2026A1515010781), Guangzhou Science and Technology Plan Project (2025A03J3929). This work was supported by Guangzhou Key Clinical Specialty (Clinical Medical Research Institute), and Guangzhou Municipal Key Discipline in Medicine (2025-2027).

## Disclosure statement

### Financial Disclosure

None.

### Non-financial Disclosure

None.

## Data availability statement

The data underlying this article will be shared on reasonable request to the corresponding author, subject to ethical and institutional restrictions.

## Notes

### Competing Interest Statement

The authors have declared no competing interest.

### Author Declarations

Ethics Committee of the Affiliated Brain Hospital of Guangzhou Medical University gave ethical approval for this work.

