## Supplementary Material for "The Circadian Disruption Index: development, validation, and responsiveness to circadian health education"

**Supplementary materials**

**Supplementary Appendix S1. Initial 22-item candidate Circadian Disruption Index**

**Circadian Disruption Index**

**Part1**

**Based on your sleep–wake schedule over the past 1 month, please answer the following questions.**

Q1 Do you usually work (or attend classes) during the daytime and sleep at night?

☐ Yes ☐ No ☐ Other: _________

Q2 What is your usual sleep period on workdays or school days?

: to :

Q3 What is your usual sleep period on free days?

: to :

Q4 How long does it usually take you to fall asleep after going to bed?

_____ minutes

Q5 What is your average duration of daytime naps (e.g., midday naps)?

_____ hours/day

Q6 How many hours of sleep per day do you need to feel fully rested?

_____ hours/day

Q7 Which chronotype best describes you?

☐ Definite morning type

☐ Moderate morning type

☐ Intermediate type

☐ Moderate evening type

☐ Definite evening type

**Part2**

**Based on your experiences over the past 1 month, please select the option that best reflects the frequency of each event.**

**Response options:**

Never (0 times/month)

Rarely (1–2 times/month)

Sometimes (1–2 times/week)

Often (3–4 times/week)

Always (daily or almost daily)

**Sleep quality and daytime functioning**

Q8 Taking ≥30 minutes to fall asleep after lights-off (without medication)

Q9 Waking up during the night and being unable to fall back asleep promptly

Q10 Perceiving insufficient sleep or poor sleep quality

Q11 Feeling sleepy during driving, work, study, or social activities

Q12 Waking up ≥2 hours later on weekends/holidays compared to weekdays

Q13 Experiencing noticeable declines in alertness or cognitive performance at certain times of the day

**Circadian rhythm disturbance–related symptoms**

Q14 Irregular sleep timing, without a consistent bedtime or wake time

Q15 Falling asleep later than desired, typically at or after 1:00 AM

Q16 Falling asleep earlier than desired, typically at or before 9:00 PM

Q17 Under free-running conditions, sleep timing progressively delays, leading to a reversal of the sleep–wake cycle

Q18 Difficulty initiating sleep or poor sleep quality due to shift work (e.g., night shifts or rotating shifts)

Q19 Difficulty initiating sleep or poor sleep quality due to transmeridian travel (jet lag)

**Behavioral and feeding rhythms**

Q20 Planned activities or meal times being disrupted by daily tasks (e.g., housework, work)

Q21 Main meals occurring at inconsistent times (variation >1 hour)

Q22 Consuming large meals or stimulants (e.g., caffeine, alcohol) before bedtime

Q23 Eating outside planned or habitual meal times (e.g., snacking, high-calorie drinks)

Q24 Engaging in vigorous activities before bedtime (e.g., exercise, outdoor play, running, wrestling)

**Light exposure and sleep environment**

Q25 First exposure to natural light occurs more than 2 hours after waking

Q26 Daily exposure to outdoor natural light is less than 1 hour

Q27 Use of electronic devices (e.g., smartphone, tablet, computer) within 1 hour before bedtime

Q28 Presence of continuous light exposure during sleep (e.g., night lights, streetlights)

Q29 Sleep being easily disturbed by light, noise, or temperature

**Supplementary Appendix S2. Final 15-item Circadian Disruption Index**

**Circadian Disruption Index**

**Part1**

**Based on your sleep–wake schedule over the past 1 month, please answer the following questions.**

Q1 Do you usually work (or attend classes) during the daytime and sleep at night?

☐ Yes ☐ No ☐ Other: _________

Q2 What is your usual sleep period on workdays or school days?

: to :

Q3 What is your usual sleep period on free days?

: to :

Q4 How long does it usually take you to fall asleep after going to bed?

_____ minutes

Q5 What is your average duration of daytime naps (e.g., midday naps)?

_____ hours/day

Q6 How many hours of sleep per day do you need to feel fully rested?

_____ hours/day

Q7 Which chronotype best describes you?

☐ Definite morning type

☐ Moderate morning type

☐ Intermediate type

☐ Moderate evening type

☐ Definite evening type

**Part2**

**Based on your experiences over the past 1 month, please select the option that best reflects the frequency of each event.**

**Response options:**

Never (0 times/month)

Rarely (1–2 times/month)

Sometimes (1–2 times/week)

Often (3–4 times/week)

Always (daily or almost daily)

**Rhythm stability and light exposur**e

Q13 Experiencing noticeable declines in alertness or cognitive performance at certain times of the day

Q15 Falling asleep later than desired, typically at or after 1:00 AM

Q21 Main meals occurring at inconsistent times (variation >1 hour)

Q25 First exposure to natural light occurs more than 2 hours after waking

Q26 Daily exposure to outdoor natural light is less than 1 hour

**Behavioral habits and diet**

Q18 Difficulty initiating sleep or poor sleep quality due to shift work (e.g., night shifts or rotating shifts)

Q20 Planned activities or meal times being disrupted by daily tasks (e.g., housework, work)

Q22 Consuming large meals or stimulants (e.g., caffeine, alcohol) before bedtime

Q23 Eating outside planned or habitual meal times (e.g., snacking, high-calorie drinks)

Q24 Engaging in vigorous activities before bedtime (e.g., exercise, outdoor play, running, wrestling)

**Sleep quality and subjective complaints**

Q8 Taking ≥30 minutes to fall asleep after lights-off (without medication)

Q9 Waking up during the night and being unable to fall back asleep promptly

Q10 Perceiving insufficient sleep or poor sleep quality

Q11 Feeling sleepy during driving, work, study, or social activities

Q29 Sleep being easily disturbed by light, noise, or temperature

**Scoring instructions**
The 15 scored items are rated from 0 to 4. Total score range: 0–60.

Factor 1 score range: 0–20.

Factor 2 score range: 0–20.

Factor 3 score range: 0–20.

Higher scores indicate greater circadian disruption.

No items are reverse scored.

**Table S1. Pre- and post-circadian health education changes in sleep-timing variables**

| **Variable (n=72)** | **Pre-Circadian**  **Health Education, mean ± SD** | **Post-Circadian**  **Health Education, mean ± SD** | **Mean change (h)** | **Wilcoxon**  ***p* value** |
| --- | --- | --- | --- | --- |
| **Workday sleep onset time** | 1.04 ± 1.02 | 0.27 ± 0.92 | -0.77 | <0.001 |
| **Free-day sleep onset time** | 1.61 ± 1.23 | 0.91 ± 0.97 | -0.70 | <0.001 |
| **Workday wake time** | 8.13 ± 0.92 | 8.06 ± 1.13 | -0.07 | 0.177 |
| **Free-day wake time** | 9.53 ± 1.09 | 9.46 ± 1.34 | -0.07 | 0.338 |
| **Workday sleep duration** | 7.09 ± 1.22 | 7.80 ± 1.15 | +0.71 | <0.001 |
| **Free-day sleep duration** | 7.93 ± 1.23 | 8.55 ± 0.99 | +0.62 | 0.001 |
| **Workday midsleep time** | 4.58 ± 0.76 | 4.17 ± 0.85 | -0.42 | <0.001 |
| **Free-day midsleep time** | 5.57 ± 0.99 | 5.19 ± 1.06 | -0.38 | 0.001 |
| **MSFsc time** | 5.13 ± 0.94 | 4.77 ± 0.90 | -0.36 | 0.002 |
| **Social jetlag** | 0.99 ± 0.73 | 1.07 ± 0.86 | +0.08 | 0.505 |

Note. All variables are presented as mean ± standard deviation in hours. Mean change was calculated as post-intervention minus pre-intervention; negative values for clock-time variables indicate earlier timing. The *p* values are from two-sided Wilcoxon signed-rank tests. SJL, social jetlag; MSFsc, sleep-corrected midsleep on free days.

**Figure S1. Pre- and post-circadian health education distributions of responses to the 15 CDI items**

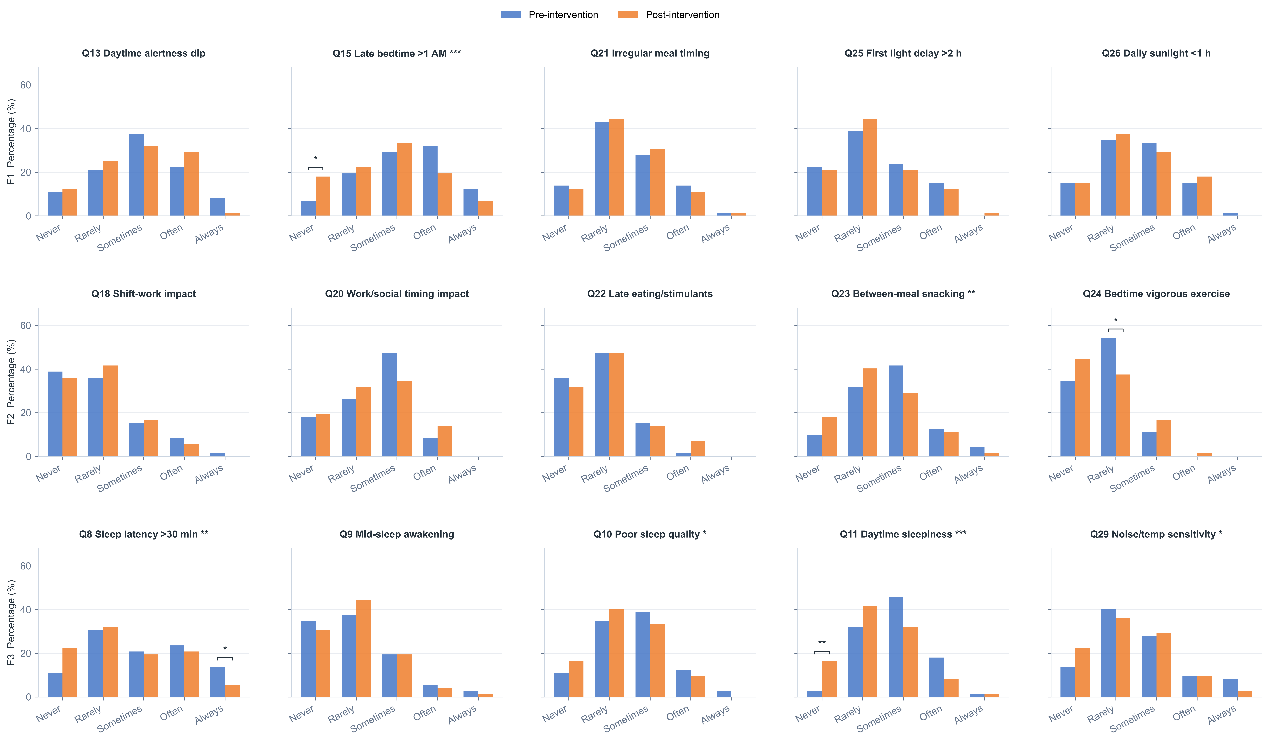

**Supplementary Figure 1. Pre- and post-Circadian Health Education distributions of responses to the 15 CDI items.** Bars show the percentage of participants selecting each response level from 0 (Never) to 4 (Always) in the paired analytic sample (N = 72). Asterisks following an item label indicate the two-sided Wilcoxon signed-rank test for the paired item score. Brackets and asterisks above a pre-post bar pair indicate the two-sided exact McNemar test for paired membership in that response category. No symbol indicates *p* ≥ 0.05; **p* < 0.05, ***p* < 0.01, and ****p* < 0.001. P values were not adjusted for multiple comparisons and should be interpreted as exploratory. CDI, Circadian Disruption Index.
